# Pre-pandemic inequalities in the burden of disease: a Scottish Burden of Disease study

**DOI:** 10.1101/2023.03.15.23286907

**Authors:** Ian Grant, Neil Chalmers, Eilidh Fletcher, Fatim Lakha, Gerry McCartney, Diane Stockton, Grant Wyper

## Abstract

**Background:** Health inequalities in Scotland are well documented, including the contribution of different causes to inequalities in mortality. Our aim was to estimate inequalities within a burden of disease framework, accounting for both premature mortality and the effects of morbidity, to understand the contribution of specific diseases to health inequalities prior to the COVID-19 pandemic.

**Methods:** Disability Adjusted Life Years (DALYs) for 70 individual causes of disease and injury were sourced from the Scottish Burden of Disease Study. Area-level deprivation was measured using the Scottish Index of Multiple Deprivation. Inequalities were measured by the range, Relative Index of Inequality, Slope Index of Inequality, and attributable DALYs were estimated by using the least deprived decile as a reference.

**Results:** The overall disease burden was double that in the most deprived areas (50,305 vs 20,955 DALYS per 100, 000), largely driven by inequalities in premature mortality. The rate in the most deprived decile was around 48% higher than the mean population rate (RII = 0.96), with 35% of DALYs attributed to differences in area-based deprivation. Many leading causes of disease burden in 2019 – heart disease, drug use disorders, lung cancer and COPD – were also the leading drivers of absolute and relative inequalities in the disease burden.

**Conclusion:** Our study evidences the extent of the stark levels of absolute and relative inequality prior to the COVID-19 pandemic. Given pre-pandemic stalling of mortality trend improvements and widening health inequalities, and the exacerbation of these caused by COVID-19, urgent policy attention is required to address this.

## Introduction

It is well evidenced that there is a clear link between levels of deprivation and life expectancy (LE) in Scotland, with those from the most deprived areas and backgrounds having significantly poorer life expectancy and general health outcomes [1, 2, 3]. Recent published data from the National Records of Scotland shows that not only have overall differences in life expectancy between the most and least deprived populations have been worsening, but the gap in the number of years of healthy life expectancy (HLE) is also widening. In terms of overall life expectancy, males in the most deprived areas have 13.5 year’s lower LE compared to their least deprived counterparts, and for females, the difference between the most and least deprived is 10.2 years [4]. Furthermore, HLE in the most deprived areas of Scotland was more than 24 years lower than in the least deprived areas for both males and females [5]. COVID-19 has further highlighted and amplified these inequalities: inequalities in COVID-19 mortality rates follow a similar social gradient to that seen for all causes of death i.e., increasing with increasing levels of deprivation, and the causes of inequalities in COVID-19 are similar to the causes of inequalities in health more generally [6,7,8].

Reliable, comparable information about the main causes of disease and injury in populations, and how these are changing and their effect on LE and HLE, is therefore a critical input for decision making, planning and targeting interventions where they are most required. Traditional sources of information about the descriptive epidemiology of diseases, injuries and risk factors are generally incomplete, fragmented and of uncertain reliability and comparability [9]. A lack of a standardized measurement framework to permit comparisons across diseases and injuries and failure to systematically evaluate data quality can impede comparative analyses of the true public health importance of various conditions and risk factors [10]. A burden of disease framework can be used to summarise the debilitating effects of morbidity and premature mortality in a population in a consistent and comparable manner. This is achieved by framing the effects of morbidity and mortality on population health loss as a function of time, in a composite measure called Disability-Adjusted Life Years (DALYs) [11]. By framing health loss in this way, DALYs combine the effects of morbidity and mortality in an equitable way and allow planners and policymakers to have a better understanding of the contribution that different diseases and injuries make to the total burden of disease [12].

Health inequalities in Scotland have been well documented, including the contribution of different causes to inequalities in mortality [1, 2, 13, 14, 15], There is, however, little information on how specific diseases contribute to these health inequalities within a standardised measurement framework, accounting for both premature mortality and the effects of morbidity. Using data from the Scottish Burden of Disease (SBoD) study, we assessed the contribution of individual disease and injuries, across a broad range of causes of morbidity and mortality, to absolute and relative health inequalities in Scotland in 2019.

## Methods

### Data

Area-level deprivation was measured using the Scottish Index of Multiple Deprivation (SIMD version 2016) [16]. The SIMD quantifies deprivation based on data zones, a geographical unit comparable to a postcode. Using pooled and weighted data from seven domains (employment, income, crime, housing, health, education and geographic access), each data zone is given a composite rank out of 6,505 data zones. The composite rank was then converted to a decile, with 1 assigned to the 10% most deprived data zones and 10 to the 10% least deprived data zones.

Estimates of the number of YLL, YLD, and DALYs for 13 broad disease groupings and 70 individual causes of disease and injury were sourced from the SBoD 2019 study [17]. YLL estimates were derived by multiplying age group-based mortality counts by the by the aspirational age-conditional life expectance from the GBD 2019 reference life table [18]. The deprivation decile was derived using the individual’s postcode of registered residence. YLD estimates were based on applying five-year age-specific rates, separately for each combination of deprivation decile for males and females, to mid-year population estimates for 2019, sourced from the National Records of Scotland [19]. As the underlying YLD rates were based on 2016 estimates, this approach assumed that any changes in the levels of YLD are entirely due to changes to the underlying demographic structure of the Scottish population.

### Analyses

All analyses were carried out at the level of age-group and SIMD decile to facilitate the calculation of age-standardised rates (ASR). ASR using mid-year population estimates were calculated directly to the 2013 European Standard Population [19,20]. ASR removes the contribution of differences in underlying population structure between SIMD deciles to facilitate like-for-like comparisons. All ASR findings are presented per 100,000 population.

We estimated inequalities using several measures. The absolute and relative range differences in ASR were estimated as the difference and ratio respectively, between the most and least deprived deciles. Additionally, inequalities were measured using the Slope Index of Inequality (SII) and Relative Index of Inequality (RII) [21,22]. This involved fitting a linear regression to the ranked area deprivation deciles and the DALY values. An equal rate across deprivation deciles would give a horizontal line with a slope of zero (SII=0), The RII was estimated by dividing the SII rate of DALYs by the overall Scottish rate of DALYs. An equal rate across deprivation deciles would give a horizontal line with a slope of zero (SII=0). Disease DALYs attributable to area deprivation were estimated by using the least deprived decile as the reference group and summing all DALYs in excess of this reference across the other nine deprivation deciles. All DALYs in excess of this reference group, were summed and expressed as a percentage of the overall disease burden [23].

Results for all inequality measurements are presented for 13 disease groups and the 20 leading individual causes of disease and injury in Scotland. All analyses were performed using R and RStudio [24,25].

## Results

### Inequalities in the disease burden

There were approximately 1,732,800 DALYs lost in Scotland in 2019 (ASR 32,093 per 100,000 population), with YLL accounting for 63.5% of total DALYs. We found evidence of marked inequalities in DALYs across areas experiencing different levels of deprivation (Figure 1). DALYs in the most deprived areas were double that of the least deprived areas (ASR: 50,305 vs. 20,955). Additionally, there were inequalities in the composition of DALYs across areas experiencing different levels of deprivation with YLL accounting for 67.8% of total DALYs in the most deprived areas, compared to 57.1% in the least deprived areas (Figure 2). The impact of this meant that the contribution of YLD to total DALYs increased with decreasing levels of deprivation, from 32.1% in the most deprived decile, rising to 42.9% in the least deprived decile. Furthermore, YLL in the most deprived decile was substantially higher (34,922 per 100,000 population) than total DALYs (20,955 per 100,000 population) in the least deprived decile (*Table 1, Supplementary file*).

**Fig 1.**
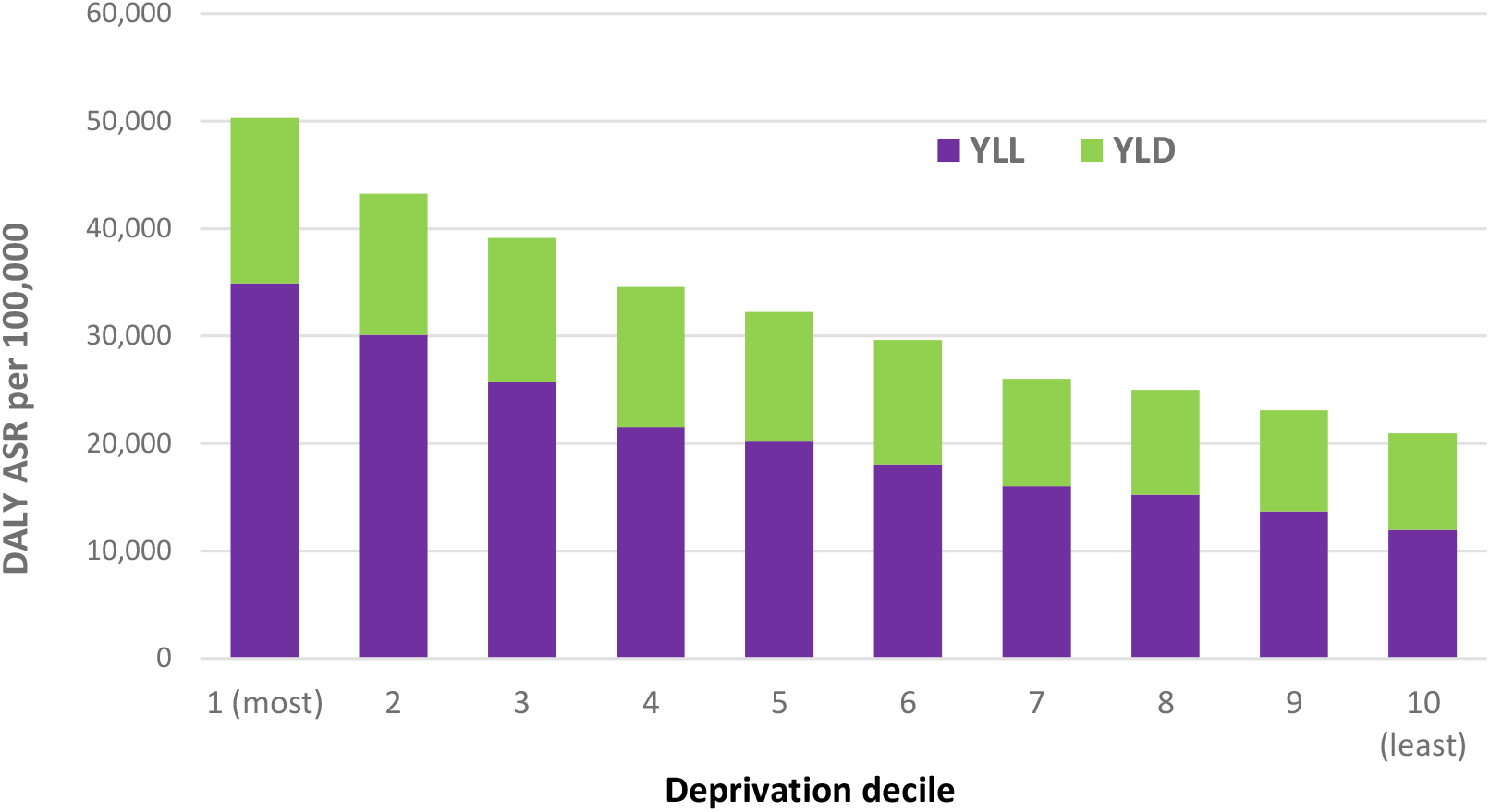
All-cause DALYs (per 100, 100) by SIMD decile, Scotland, 2019.

**Fig 2.**
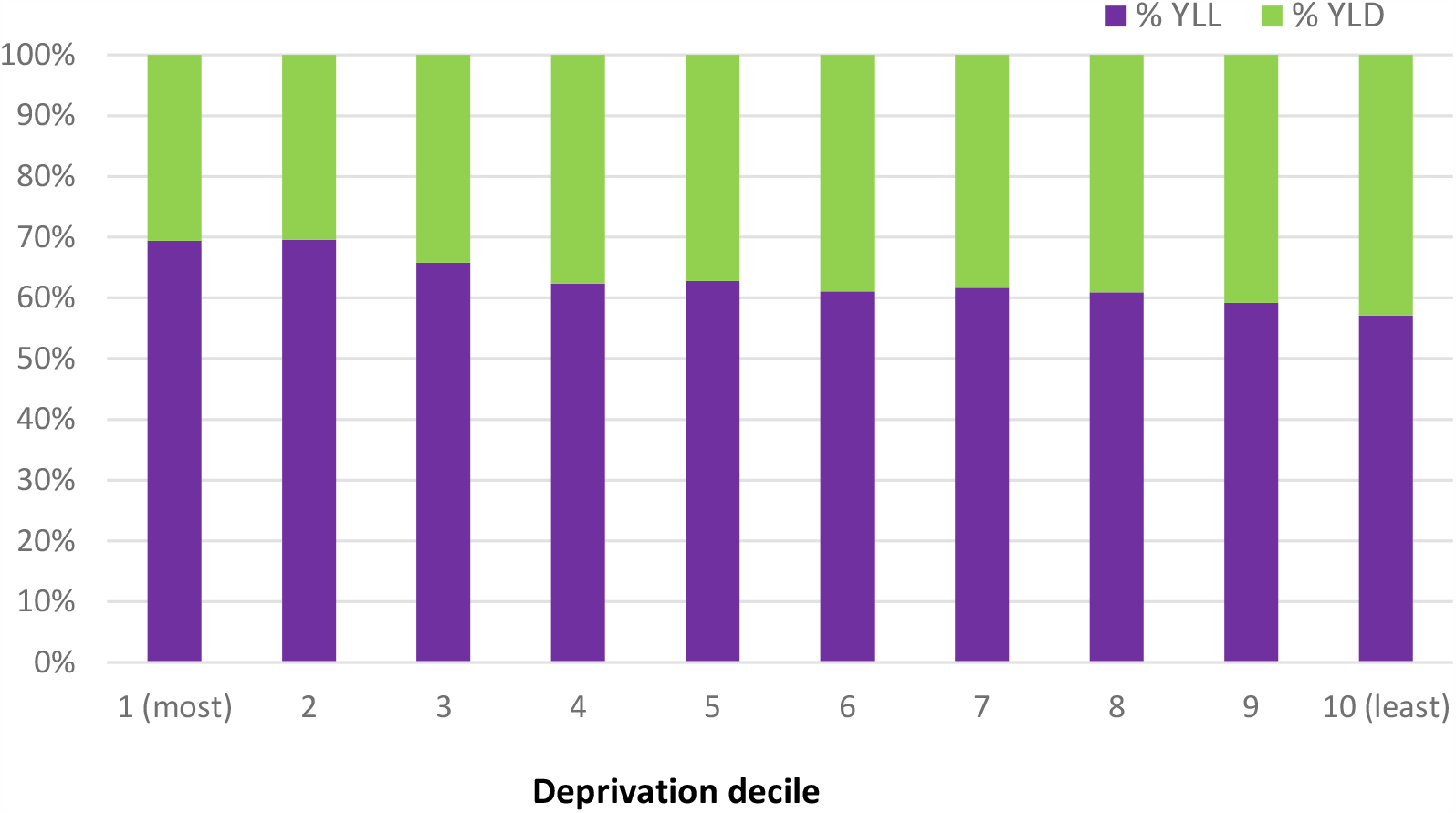
Proportion of YLL and YLD by SIMD decile, Scotland 2019.

**Table 1.** Inequalities in the broad causes of DALYs, per 100, 00 population, Scotland, 2019

| Disease group | ASR of DALYs per 100,000 | Absolute rate difference between most and least deciles | Relative rate difference between most and least deciles | Slope Index of Inequality | Relative Index of Inequality | Attributable DALYs (%) |
| --- | --- | --- | --- | --- | --- | --- |
| Cancers | 6,460 | 4,780 | 2.0 | 4,831 | 0.75 | 27.6 |
| Cardiovascular diseases | 5,416 | 4,583 | 2.3 | 5,206 | 0.96 | 36.3 |
| Neurological disorders | 3,534 | 1,009 | 1.4 | 1,069 | 0.30 | 22.3 |
| Mental health disorders | 2,494 | 2,464 | 2.6 | 2,622 | 1.05 | 38.3 |
| Musculoskeletal disorders | 2,263 | 399 | 1.2 | 530 | 0.23 | 13.7 |
| Substance use disorders | 2,250 | 6,837 | 20.5 | 6,334 | 2.82 | 84.5 |
| Chronic respiratory diseases | 1,871 | 2,627 | 4.0 | 2,788 | 1.49 | 54.0 |
| Injuries | 1,646 | 1,980 | 3.1 | 2,061 | 1.25 | 43.5 |
| Other non-communicable diseases | 1,506 | 826 | 1.7 | 922 | 0.61 | 24.2 |
| Digestive diseases | 1,503 | 1953 | 3.5 | 2,118 | 1.41 | 49.7 |
| Diabetes and chronic kidney diseases | 1,133 | 1,028 | 2.5 | 1,108 | 0.98 | 38.3 |
| Communicable, maternal, neonatal, and nutritional diseases | 1,119 | 1,069 | 2.5 | 1,092 | 0.98 | 38.3 |
| Sensory conditions | 815 | -204 | -0.8 | -169 | -0.21 | -13.8 |
| All causes | 32,093 | 29,350 | 2.4 | 30,512 | 0.95 | 35.3 |

### Absolute and relative inequalities in all cause DALYs

The absolute difference in all-cause DALYs between the most and least deprived areas in Scotland was approximately 29,350 per 100,000 population (Table 1). The relative difference in DALYs between the most and least deprived areas was 2.4, indicating that DALYs were 2.4 times higher in our most, compared to least, deprived areas. The RII was 0.95, which when multiplied by 0.5 and expressed as a percentage means that the rate in the most deprived areas was 47.5% higher than the mean rate of DALYs in the Scottish population. The difference between the most and least deprived areas, measured by SII was 30,502 DALYs per 100,000 population. DALYs attributable to differences in area deprivation accounted for 35% of total DALYs.

### Absolute and relative inequalities in disease groupings

In terms of absolute rate differences of DALYs, between the most and deprived areas, substance use disorders had the largest difference (6,837 DALYs per 100,000 population), followed by cancers (4,783 DALYs per 100,000 population) and cardiovascular diseases (4,583 DALYs per 100,000 population) (Table 1). For all others disease groups, except sensory conditions, there were positive absolute rate differences, indicating that the rate of DALYs were higher in the most, compared to least deprived areas.

Relative rate differences between the most and least deprived areas were also evident. In over half (9 out of 17) of the disease groups, the relative rate difference was ≥2.0. Substance use disorders had the highest relative rate difference (20.5), followed by chronic respiratory diseases (4.0) and digestive diseases (3.5). These three disease groups also had the highest RII values indicating that the rate in most deprived communities was higher than the mean rate of population: substance use disorders (RII = 2.82); chronic respiratory diseases (RII=1.49), digestive diseases (RII=1.41).

The disease groups with the largest estimates of DALYs attributable to inequalities in area deprivation were: substance use disorders (84.5%); chronic respiratory diseases (54.0%) and digestive diseases (49.7%). Conversely, we found a negative association for DALYs inequality attributable DALYs for sensory conditions (−13.8%). The estimates of inequality attributable DALYs for all other disease groups ranged from 13.7% for musculoskeletal disorders to 43.5% for injuries.

### Absolute and relative inequalities in individual causes

Of the 20 leading causes of diseases burden, drug use disorders had the largest absolute rate difference (5,452 DALYs per 100,000 population) between the most and least deprived areas, followed by ischaemic heart disease (3,027 DALYs per 100,000 population), lung cancer (2,475 DALYs per 100,000 population) and chronic obstructive pulmonary disorder (2,358 DALYs per 100,000 population) (Table 2). For most of the remaining causes, there were positive absolute rate differences in DALYs, indicating DALYs were higher in the most compared to least deprived areas. The only exception to this was headache disorders, which had a negligible negative estimate of -15 DALYs per 100,000 population.

**Table 2.** Inequalities in the 20 leading causes of DALYs, Scotland, 2019

| Leading causes | ASR of DALYs per 100,000 | Absolute rate difference between most and least deciles | Relative rate difference between most and least deciles | Slope Index of Inequality | Relative Index of Inequality | Attributable DALYs (%) |
| --- | --- | --- | --- | --- | --- | --- |
| Ischaemic heart disease | 2,572 | 3,027 | 3.2 | 3,165 | 1.23 | 46.2 |
| Alzheimer's disease and other dementias | 1,764 | 492 | 1.4 | 576 | 0.33 | 25.0 |
| Drug use disorders | 1,673 | 5,452 | 28.3 | 5,003 | 2.99 | 88.0 |
| Tracheal, bronchus, and lung cancer | 1,596 | 2,475 | 4.3 | 2,518 | 1.58 | 54.9 |
| Cerebrovascular disease | 1,450 | 790 | 1.8 | 1,022 | 0.70 | 30.7 |
| Chronic obstructive pulmonary disease | 1,293 | 2,358 | 6.4 | 2,442 | 1.89 | 67.6 |
| Low back and neck pain | 1,269 | 355 | 1.3 | 430 | 0.34 | 17.5 |
| Depression | 1,243 | 1,212 | 2.6 | 1,282 | 1.03 | 38.2 |
| Other cancers | 1183 | 955 | 2.1 | 865 | 0.73 | 24.8 |
| Headache disorders | 982 | -19 | 1.0 | 20 | 0.02 | 15.0 |
| Other cardiovascular and circulatory diseases | 908 | 574 | 1.8 | 738 | 0.81 | 30.2 |
| Anxiety disorders | 852 | 809 | 2.5 | 861 | 1.01 | 37.9 |
| Self-harm and suicide | 789 | 1,315 | 4.9 | 1,279 | 1.62 | 56.8 |
| Colon and rectum cancer | 752 | 226 | 1.3 | 219 | 0.29 | 21.4 |
| Diabetes mellitus | 744 | 688 | 2.5 | 760 | 1.02 | 39.7 |
| Lower respiratory infections | 699 | 670 | 2.5 | 717 | 1.03 | 37.9 |
| Cirrhosis and other chronic liver diseases | 626 | 1,107 | 5.4 | 1,164 | 1.86 | 62.6 |
| Alcohol use disorders | 577 | 1,385 | 10.2 | 1,332 | 2.31 | 74.4 |
| Breast cancer | 555 | 84 | 1.2 | 77 | 0.14 | 8.0 |
| Other digestive diseases | 431 | 465 | 3.0 | 502 | 1.17 | 47.9 |
| All cause DALY | 32,093 | 29,350 | 2.4 | 30,512 | 0.96 | 35.3 |

Relative rate differences between the most and least deprived areas were also evident across most of the leading individual causes of disease burden. Drug use disorders had the highest relative rate difference (28.3) then followed by alcohol use disorders (10.2), chronic obstructive pulmonary disorder (6.4), cirrhosis and other chronic liver diseases (5.4) and self-harm and suicide (4.9).

The diseases with the highest relative rate differences also had some of the highest RII values, indicating that the rate in most deprived communities was substantially higher than the mean rate of population (Scotland-level RII=0.96) for these diseases: substance use disorders (RII=2.99), alcohol use disorders (RII=2.31), chronic obstructive pulmonary disorder (RII=1.89), cirrhosis and other chronic liver diseases (RII=1.86), self-harm and suicide (RII=1.6) and lung cancer (RII=1.6). Notably, several conditions out with the leading causes of disease burden in Scotland had high RII values in the most deprived decile compared to mean rate of population, including schizophrenia (RII=5.5) and epilepsy (RII=4.7) (*Table 2, Supplementary file*).

The estimate of all-cause inequality attributable DALYs was 35.3%, however this varied considerably by individual cause (Table 2). Of the twenty leading causes of disease burden, DALYs attributable to inequalities in area deprivation ranged from 88.0% for drug use disorders, 74.4% for alcohol use disorders DALYs and 67.6 % for chronic obstructive pulmonary disorder, to 17.5% for low back and neck pain and 10% of headache disorder DALYs.

## Discussion

Our study evidences the extent of stark levels of absolute and relative inequality, using range and distributional measures, in Scotland in 2019, prior to the COVID-19 pandemic. The overall disease burden was double that in the most deprived areas in Scotland, largely driven by inequalities in mortality. We found that the gap in the burden of disease is so wide that the fatal burden of disease (YLL) in the most deprived areas was substantially higher than the total burden of disease (DALYs) in the least deprived areas. We already know from life expectancy figures that people in the most deprived communities die at younger age and spend more than a third of their lives in poor health compared to the least deprived communities [4,5]. Our findings further strengthen our understanding of the cause specific diseases driving the interaction between stalling mortality improvements, HLE and health inequalities [26,27].

Many of the leading causes of disease burden in 2019 – heart disease, drug use disorders, lung cancer and chronic obstructive pulmonary disease – were also the leading drivers of absolute and relative inequalities in the disease burden. Causes such as: alcohol, and drug use disorders; liver cirrhosis; chronic obstructive pulmonary disease; lung cancer; and, self-harm and suicide, exhibited the largest relative inequalities in disease burden which is consistent with results published elsewhere in the literature [2, 13, 15]. On the other hand, there were health conditions where there were smaller, to no, inequalities, for example: sensory conditions; headaches; and some musculoskeletal conditions. Triangulating these results with estimates of inequality-attributable DALY estimates for COVID-19, indicate that although COVID-19 infection has likely increased all-cause inequalities, there are several other causes of disease which have a much larger impact in driving levels of all-cause inequality in the burden of disease [8].

The COVID-19 pandemic has caused almost unprecedented change across health, education and the economy in Scotland and across the world. COVID-19 has been shown to have a substantial impact on the population health in Scotland, however this impact was not shared equally across areas experiencing different levels of socio-economic deprivation; in Scotland, the marked inequalities in COVID-19 YLL reported in 2020 were further exacerbated in 2021, to the extent that approximately half of COVID-19 YLL was attributable to inequalities in area deprivation. This widening of inequalities in 2021 was confirmed across all measures of absolute and relative inequality [7, 8]. Furthermore, health problems that existed before the COVID pandemic, have not gone away, and in many cases have been exacerbated, and unless future harm is prevented, this trend will continue. Integrating this impact, quantitively, is difficult, and trends may not be comparable due to the major disruptions, and changes in access to, and delivery of, services, and the competition between causes of mortality [28]. Prior to the pandemic, improvements in life expectancy and healthy life expectancy in Scotland had stalled since 2012, with a slowdown in the overall progress of reducing mortality and widening of socioeconomic inequalities in mortality [5, 29,30, 35]. The results presented in this paper are therefore likely to be representative of wider pre pandemic trends given that YLD does not change quickly over time and that YLL estimates ultimately drive changes in the overall DALYs. Our estimates help us to understand the state of pre-pandemic inequalities, providing important baseline positions that can be triangulated with robust, and locally relevant, emerging evidence, to assess the likely impact of the pandemic, and other public health crisis such as the current cost-of-living crisis, on non-COVID and COVID-related health inequalities.

### Strengths and Limitations

This study has several important strengths. Firstly, this study assesses the impact of inequalities using DALYs for over 70 individual diseases, conditions and injuries. DALYs provide a composite, internally consistent measure of population health loss which can be used to evaluate the proportionate burden of different diseases and injuries and compare population health by geographic region and over time. By combining information on fatal and non-fatal burden, BOD studies allow planners and policymakers to have a better understanding of the contribution that different diseases and injuries make to the total burden of disease and how this burden varies by levels of deprivation. This in turn supports decisions about where prevention and service activity should be focused to address health inequalities in Scotland.

YLD estimates are derived from surveys, and routine administrative data and as such may not be fully representative of the underlying burden by deprivation decile, for example administrative data are reflective of demand rather than need. This is also relevant for severity distributions, which are internationally derived with cancers being the main exception. In terms of impact of study results by deprivation decile, on the absolute scale this is most relevant for the causes with the largest YLD estimates, or those where YLD is the main contributor to disease-specific DALYs. For those causes which have a larger YLL component, it’s plausible that any biases in YLD from unmet need or survey-specific bases will have a smaller impact on the DALY estimate if this unmet need causes disease-specific excess mortality.

Our measure of deprivation is area-based rather than individual measure and is therefore likely to misclassify many individuals into categories that do not reflect their individual experiences [32]. Although the SIMD includes health indicators in its range of domains (thereby raising the theoretical possibility of reverse causality whereby people are ordered by the health outcome rather than socioeconomic deprivation), the employment-income deprivation index (i.e., excluding the health measures) is very highly correlated with the overall SIMD index and so this is unlikely to have changed the results [33,34]. Furthermore, The SII and RII have the advantage that they are based on data about the whole population, rather than just the extremes, and so take into account inequalities across the entire distribution of inequality. They do, however, require a reasonably linear relationship between outcomes and exposures [31].

## Conclusion

This study demonstrates that even before the COVID-19 pandemic there was a steep stepwise gradient in the burden of disease experienced across the Scottish population. This was evident for YLL and YLD, and almost every specific cause of death. By exploring cause-specific inequalities within a standardized measurement framework, to permit equitable comparisons across diseases and injuries, we confirm the stark gap between the most and least deprived areas in Scotland in 2019 prior to the COVID-19 pandemic and show that many of the leading causes of disease burden in 2019 were also the leading drivers of absolute and relative health inequalities in the disease burden.

Given the pre-pandemic stalling of mortality trend improvements and widening health inequalities, and the exacerbation of these caused by COVID-19, urgent policy attention is required to reduce the burden of fatal and non-fatal conditions in Scotland.

## Data Availability

All data produced in the present study are available upon reasonable request to the authors

