## Supplementary Tables for "Pre-pandemic inequalities in the burden of disease: a Scottish Burden of Disease study"

**Supplementary Table 1. All-cause DALYs (European age standardised rate per 100, 000) by Scottish Index of Multiple Deprivation Decile, Scotland, 2019**

|  | **DALY** | | **YLL** | | **YLD** | |  |
| --- | --- | --- | --- | --- | --- | --- | --- |
| **SIMD deciles** | **Number** | **EASR**  **per 100,000** | **Number** | **EASR**  **per 100,000** | **Number** | **EASR per 100,000** | **% YLD** |
| **1 (most deprived)** | 244,777 | 50,305 | 166,088 | 34,922 | 78,690 | 15,383 | 0.31 |
| **2** | 218,789 | 43,256 | 150,192 | 30,091 | 68,596 | 13,165 | 0.30 |
| **3** | 206,329 | 39,132 | 134,827 | 25,736 | 71,502 | 13,397 | 0.34 |
| **4** | 187,256 | 34,585 | 116,492 | 21,561 | 70,764 | 13,024 | 0.38 |
| **5** | 180,781 | 32,235 | 114,606 | 20,247 | 66,176 | 11,989 | 0.37 |
| **6** | 167,651 | 29,608 | 103,165 | 18,080 | 64,486 | 11,528 | 0.39 |
| **7** | 146,328 | 26,026 | 90,520 | 16,046 | 55,808 | 9,980 | 0.38 |
| **8** | 139,140 | 24,975 | 84,510 | 15,208 | 54,630 | 9,767 | 0.39 |
| **9** | 127,178 | 23,106 | 75,189 | 13,669 | 51,989 | 9,436 | 0.41 |
| **10 (least deprived)** | 114,572 | 20,955 | 65,407 | 11,962 | 49,165 | 8,994 | 0.43 |

**Supplementary Table 2. DALY inequalities in the Scottish Burden of Disease Study, 2019**

| **Cause** | **DALY ASR per 100,000** | **Absolute rate difference between most and least deciles** | **Relative rate difference** **between most and least deciles** | **SII per 100,000** | **RII** | **Attributable DALYs (%)** |
| --- | --- | --- | --- | --- | --- | --- |
| Ischaemic heart disease | 2572.2 | 3,027 | 3.2 | 3,165 | 1.2 | 46.2 |
| Alzheimer's disease and other dementias | 1763.9 | 492 | 1.4 | 576 | 0.3 | 25.0 |
| Drug use disorders | 1673.1 | 5,452 | 28.3 | 5,003 | 3.0 | 88.0 |
| Lung cancer | 1595.9 | 2,475 | 4.3 | 2,518 | 1.6 | 54.9 |
| Cerebrovascular disease | 1449.7 | 790 | 1.8 | 1,022 | 0.7 | 30.7 |
| Chronic obstructive pulmonary disease | 1292.9 | 2,358 | 6.4 | 2,442 | 1.9 | 67.6 |
| Low back and neck pain | 1269.2 | 355 | 1.3 | 430 | 0.3 | 17.5 |
| Depression | 1243.3 | 1,212 | 2.6 | 1,282 | 1.0 | 38.2 |
| Other cancers | 1182.9 | 955 | 2.1 | 865 | 0.7 | 24.8 |
| Headache disorders | 981.6 | -19 | 1.0 | 20 | 0.0 | 15.0 |
| Other cardiovascular and circulatory diseases | 907.7 | 574 | 1.8 | 738 | 0.8 | 30.2 |
| Anxiety disorders | 852.1 | 809 | 2.5 | 861 | 1.0 | 37.9 |
| Self-harm and interpersonal violence | 789.1 | 1,315 | 4.9 | 1,279 | 1.6 | 56.8 |
| Colorectal cancer | 752.1 | 226 | 1.3 | 219 | 0.3 | 21.4 |
| Diabetes mellitus | 743.7 | 688 | 2.5 | 760 | 1.0 | 39.7 |
| Lower respiratory infections | 698.8 | 670 | 2.5 | 717 | 1.0 | 37.9 |
| Cirrhosis and other chronic liver diseases | 625.8 | 1,107 | 5.4 | 1,164 | 1.9 | 62.6 |
| Alcohol use disorders | 577.4 | 1,385 | 10.2 | 1,332 | 2.3 | 74.4 |
| Breast cancer | 555.2 | 84 | 1.2 | 77 | 0.1 | 8.0 |
| Other digestive diseases | 431.0 | 465 | 3.0 | 502 | 1.2 | 47.9 |
| Other musculoskeletal disorders | 426.4 | -17 | 1.0 | -10 | -0.02 | 15.7 |
| Falls | 425.4 | 319 | 2.0 | 374 | 0.9 | 33.4 |
| Osteoarthritis | 389.2 | 66 | 1.2 | 82 | 0.2 | 13.1 |
| Chronic kidney disease | 386.6 | 329 | 2.3 | 341 | 0.9 | 35.3 |
| Atrial fibrillation and flutter | 380.2 | 88 | 1.3 | 141 | 0.4 | 19.0 |
| Skin and subcutaneous diseases | 369.0 | 55 | 1.1 | 60 | 0.2 | 10.4 |
| Prostate cancer | 354.0 | 57 | 1.2 | 77 | 0.2 | 19.1 |
| Oesophageal cancer | 337.9 | 293 | 2.2 | 217 | 0.6 | 31.0 |
| Pancreatic cancer | 331.4 | 52 | 1.2 | 114 | 0.3 | 17.6 |
| Age-related and other hearing loss | 318.9 | -105 | 0.7 | -113 | -0.4 | 22.8 |
| Endocrine, metabolic, blood, and immune disorders | 306.7 | 374 | 3.6 | 322 | 1.1 | 52.7 |
| Asthma | 296.0 | 31 | 1.1 | 49 | 0.2 | 20.8 |
| Other chronic respiratory diseases | 282.0 | 238 | 2.1 | 297 | 1.1 | 35.7 |
| Epilepsy | 271.2 | 436 | 4.7 | 401 | 1.5 | 57.0 |
| Congenital birth defects | 253.9 | 155 | 1.9 | 180 | 0.7 | 34.0 |
| Other sense organ diseases | 251.9 | -78 | 0.7 | -72 | -0.3 | 24.3 |
| Blindness and vision loss | 244.2 | -21 | 0.9 | 16 | 0.1 | 12.3 |
| Urinary diseases and male infertility | 233.7 | 88 | 1.5 | 151 | 0.6 | 29.6 |
| Liver cancer | 231.3 | 167 | 1.9 | 245 | 1.1 | 40.6 |
| Schizophrenia | 224.1 | 401 | 5.5 | 416 | 1.9 | 61.0 |
| Other unintentional injuries | 222.7 | 198 | 2.3 | 230 | 1.0 | 50.0 |
| Oral disorders | 203.0 | 62 | 1.3 | 83 | 0.4 | 19.6 |
| Other neurological disorders | 199.7 | 81 | 1.6 | 45 | 0.2 | 28.5 |
| Gynecological diseases | 188.1 | 24 | 1.1 | 34 | 0.2 | 11.3 |
| Parkinson's disease | 182.1 | -14 | 0.9 | -24 | -0.1 | 17.8 |
| Bladder cancer | 181.2 | 160 | 2.1 | 141 | 0.8 | 25.7 |
| Kidney cancer | 180.2 | 118 | 2.2 | 93 | 0.5 | 45.7 |
| Rheumatoid arthritis | 178.3 | -5 | 1.0 | 28 | 0.2 | 21.3 |
| Other mental health disorders | 174.2 | 43 | 1.3 | 64 | 0.4 | 17.2 |
| Ovarian cancer | 166.5 | 54 | 1.4 | 62 | 0.4 | 28.0 |
| Non-Hodgkin's lymphoma | 165.3 | 54 | 1.5 | 80 | 0.5 | 32.1 |
| Stomach cancer | 164.3 | 135 | 2.0 | 148 | 0.9 | 41.0 |
| Gallbladder and biliary diseases | 156.6 | 127 | 2.4 | 127 | 0.8 | 42.8 |
| Transport injuries | 154.0 | 79 | 1.8 | 112 | 0.7 | 38.0 |
| Leukaemia | 152.2 | -67 | 0.6 | -27 | -0.2 | 33.6 |
| Diarrhoea and other common infections | 141.3 | 111 | 2.2 | 105 | 0.7 | 36.2 |
| Multiple sclerosis | 135.2 | 32 | 1.3 | 51 | 0.4 | 17.2 |
| Neonatal disorders | 118.4 | 88 | 2.5 | 107 | 0.9 | 50.8 |
| Inflammatory bowel disease | 116.2 | 28 | 1.3 | 33 | 0.3 | 16.3 |
| Multiple myeloma | 110.0 | 17 | 1.2 | 1 | 0.0 | 21.9 |
| Peripheral vascular disease | 106.0 | 105 | 2.8 | 140 | 1.3 | 51.0 |
| Peptic ulcer disease | 90.1 | 134 | 3.7 | 165 | 1.8 | 58.1 |
| Nutritional deficiencies | 83.7 | 74 | 2.4 | 56 | 0.7 | 36.4 |
| Pancreatitis | 82.9 | 92 | 3.0 | 126 | 1.5 | 46.5 |
| Unknown cause of injury | 54.8 | 68 | 3.0 | 65 | 1.2 | 38.9 |
| Sudden infant death syndrome | 34.9 | 68 | - | 93 | 2.7 | - |
| Other communicable, maternal, neonatal and nutritional diseases | 31.6 | 7 | 1.3 | 12 | 0.4 | 25.9 |
| HIV/AIDS and tuberculosis | 29.5 | 96 | 12.2 | 73 | 2.5 | 77.2 |
| Maternal disorders | 15.5 | 22 | 2.6 | 23 | 1.5 | 57.5 |
| Other diabetes and chronic kidney disorders | 2.9 | 11 | 525.8 | 7 | 2.4 | 99.7 |
